# A population-level analysis of the protective effects of androgen deprivation therapy against COVID-19 disease incidence and severity

**DOI:** 10.1101/2021.05.10.21255146

**Authors:** Kyung Min Lee, Kent Heberer, Anthony Gao, Daniel J Becker, Stacy Loeb, Danil V. Makarov, Barbara Gulanski, Scott L DuVall, Mihaela Aslan, Jennifer Lee, Richard Hauger, Mei-Chiung Shih, Julie Lynch, Matthew Rettig

## Abstract

**Importance:** The incidence and severity of coronavirus disease 19 (COVID-19) is higher in men. Sex hormones potentially offer one explanation for differences by sex.

**Objective:** To determine whether men exposed to androgen deprivation therapy (ADT) have lower incidence and severity of COVID-19.

**Design:** We conducted an observational study of male Veterans treated in the Veterans Health Administration from February 15th to July 15th, 2020. We developed a propensity score model to predict the likelihood to undergo Severe Acute Respiratory Syndrome Coronavirus 2 (SARS-CoV-2) testing. We performed multivariable logistic regression modeling adjusted with inverse probability weighting to examine the relationship between ADT and COVID-19 incidence. We conducted logistic regression analysis among COVID-19 patients to test the association between ADT and COVID-19 severity.

**Setting:** The U.S. Department of Veterans Affairs

**Participants:** The study sample consisted of 6,250,417 male Veterans who were alive as of February 15, 2020.

**Exposure:** Exposure to ADT was defined as having any prescription for a luteinizing hormone releasing hormone analogue or an antiandrogen in the six months prior to the index date.

**Main Outcomes and Measures:** To assess incidence, we used a binary variable indicating any positive reverse transcriptase polymerase chain reaction SARS-CoV-2 test result through July 15, 2020. To measure severity, we constructed a binary variable indicating whether a patient was admitted to the intensive care unit, placed on mechanical ventilation, or dead in the 60 days following a positive test up to July 15, 2020.

**Results:** We identified 246,087 patients who had been tested for SARS-CoV-2, of whom 3,057 were exposed to ADT, and 36,096 patients with cancer and no ADT exposure. Of these, 295 ADT patients and 2,427 other cancer patients had COVID-19 illness. In the primary, propensity-weighted comparison of ADT patients to cancer patients not on ADT, ADT was associated with decreased likelihood of testing positive for SARS-CoV-2 (adjusted OR, 0.88 [95% CI, 0.81-0.95]; p=0.001). ADT was associated with fewer severe COVID-19 outcomes (OR 0.72 [95% CI 0.53-0.96]; p=0.03).

**Conclusions and Relevance:** ADT is associated with reduced incidence and severity of COVID-19 amongst male Veterans. Repurposing of drugs that modulate androgen production and/or action may represent viable potential treatments for COVID-19.

**KEY POINTS:** *Question:* Does androgen deprivation therapy (ADT) lower incidence and severity of COVID-19?

*Findings:* In this observational study of male Veterans treated in the Veterans Healthcare System, ADT was associated with decreased likelihood of testing positive for SARS-CoV-2 (adjusted OR, 0.88 [95% CI, 0.81-0.95]; p=0.001). ADT was also associated with fewer severe COVID-19 outcomes (OR 0.72 [95% CI 0.53-0.96]; p=0.03).

*Meaning:* The use of androgen deprivation therapy may be protective against SARS-CoV-2 infection and modulate severity of COVID-19 outcomes.

## INTRODUCTION

Severe acute respiratory syndrome coronavirus 2 (SARS-CoV-2) infection and coronavirus disease 2019 (COVID-19) have rapidly disseminated across the globe and caused more than one million deaths.^1^ The incidence and severity of COVID-19 have been related to multiple factors, including age, comorbid conditions, immunosuppression, smoking, race, and sex.^2–4^ Whereas men and women manifest a similar incidence of COVID-19, male sex is a risk factor for more severe illness. The severity of COVID-19 as indicated by admission to an intensive care unit (ICU), requirement for mechanical ventilation, or mortality is approximately two-fold higher in men.^5,6^ This sex disparity has been observed in Asia, North America, Europe, and across individual countries on each of the continents.

Explanations for sex differences in outcome include difference in health behaviors (e.g. smoking history), incidence of comorbidities such as lung and heart disease, and biology, including variations in sex hormones. Specifically, androgens including testosterone and dihydrotestosterone may worsen COVID-19 severity through two non-mutually exclusive physiologic effects. First, androgens are known to suppress both innate and adaptive immunity, which may increase susceptibility to the SARS-CoV-2 virus and disease severity.^7,8^ Second, androgens induce the expression of the two plasma membrane proteins, transmembrane protease serine 2 (TMPRSS2) via an androgen response element in the promoter and angiotensin converting enzyme 2 (ACE2), which are required for entry of SARS-CoV-2 into epithelial cells.^9–12^ Co-expression of the androgen receptor (AR), TMPRSS2, and ACE2 has been observed in human pulmonary epithelial cells, alveolar type 2 pneumocytes, and nasal mucosal cells which are critical sites for SARS-CoV-2 infection and disease severity.^12,13^ Androgen-driven overexpression of these viral co-receptors may augment cellular entry and replication of SARS-CoV-2, thereby mediating increased infection rate and COVID-19 severity in men. Furthermore, men who are carriers of TMPRSS2 gene mutations, which potentiate androgen-induced cellular expression of TMPRSS2, may be especially at risk for COVID-19 morbidity and mortality.^14,15^ TMPRSS2 gene variants has also been found to confer a two-fold increase in H1N1 influenza severity supporting the hypothesis that androgen regulation of TMPRSS2 may be a critical host factor for respiratory disease.^16^

These observations suggest that suppression of androgen levels may have a salutary impact on COVID-19 illness. Two retrospective studies that evaluated the impact of androgen deprivation therapy (ADT) on COVID-19 illness yielded conflicting results. A study from Italy reported a reduced incidence of COVID-19 among patients with prostate cancer on ADT, whereas a study of patients in the United States (U.S.) failed to detect a protective effect of ADT on COVID-19 incidence.^17,18^ Our objective was to perform a nationwide, population-based study of the relationship of ADT to the incidence and outcomes of COVID-19. Using data from the U.S. Veterans Health Administration (VHA), we report the largest study to date evaluating the hypothesis that androgen deprivation therapy is associated with a reduced incidence and severity of COVID-19.

## METHODS

### Data sources

We used the Corporate Data Warehouse (CDW) of the VHA, a national data repository that provides access to the electronic health records of all individuals who received care in the VHA. In addition, we drew from the VA COVID-19 Shared Data Resource (CSDR), a newly developed data domain that consists of a wide range of information related to COVID-19 available for all patients who received a COVID-19 laboratory test within VHA or whose positive test result outside VHA was recorded in VHA clinical notes.^19^ This study was approved by the VA Central Institutional Review Board.

### Study sample

The study sample consisted of all male Veterans who were alive as of February 15, 2020. The earliest testing date reported in the CSDR is February 16, 2020. Therefore, we considered anyone alive as of February 15, 2020 as having been eligible to be tested for SARS-CoV-2. From this base sample, we assembled two sub-samples to examine incidence of COVID-19 and severity of COVID-19. For incidence analysis, we derived a sample consisting of patients who underwent SARS-CoV-2 testing matched to patients who did not undergo SARS-CoV-2 testing on age, race, and VHA facility. Separately, we used an unmatched sample of patients who tested positive for SARS-CoV-2 to investigate ADT use and COVID-19 severity. The index date for the incidence analysis and the severity analysis was February 15, 2020 and the date of first positive SARS-CoV-2 test result, respectively.

### Exposure

Exposure to androgen deprivation therapy (ADT) was defined as having any prescription for a luteinizing hormone releasing hormone analogue (LHRH) (buserelin, degarelix, goserelin, historelin, leuprolide, or triptorelin) or an antiandrogen (abiraterone, apalutamide, bicalutamide, cyproterone, darolutamide, enzalutamide, flutamide, ketoconazole, or nilutamide) in the six months prior to the index date. A small number of patients were prescribed only antiandrogens. These patients were presumed to be receiving their LHRH analogues outside the VHA because it is highly unlikely to be prescribed antiandrogens as a monotherapy.

### Endpoints

We investigated the relationship between ADT use and COVID-19 incidence and severity. To assess incidence, we used a binary variable indicating any positive reverse transcriptase polymerase chain reaction (RT-PCR) SARS-CoV-2 test result through July 15, 2020. To measure severity, we constructed a binary variable indicating whether a patient was admitted to the intensive care unit (ICU), placed on mechanical ventilation, or dead in the 60 days following a positive test up to July 15, 2020. Death was attributed to COVID-19 if it occurred within 60 days of a positive RT-PCR test for SARS-CoV-2. The primary comparison for both incidence and severity was patients with ADT exposure (ADT patients) versus patients with non-prostate cancer who had no ADT exposure (non-ADT, other cancer patients). We selected this as the primary comparison because the incidence and severity of COVID-19 is adversely affected by a cancer diagnosis and virtually all ADT patients have high risk, recurrent, locally advanced, or metastatic prostate cancer. Secondary comparisons included all patients exposed to ADT versus all who were not, and patients with prostate cancer with versus without ADT exposure.

### Covariates

Age, race, marital status, body mass index, smoking status, the Charlson Comorbidity Index categories^20^ (excluding localized and metastatic solid tumors) in the prior two years, use of selected antihypertensives—angiotensin-converting enzyme inhibitors (ACEs), angiotensin II receptor blockers (ARBs), and spironolactone—in the prior six months, VHA utilization in the prior year as measured by total numbers of visits and hospital days, and the number of days to testing or first SARS-CoV-2 positivity were assessed at the index date. Lastly, the VHA facility of COVID-19 testing was included as a fixed effect in the COVID-19 incidence models and as a random effect in the COVID-19 severity models.

### Statistical analysis

Multivariate logistic regression models were used to determine the association of ADT exposure with SARS-CoV-2 positivity, weighted by the inverse of the predicted probabilities of being tested. Due to the limited availability of SARS-CoV-2 tests, receipt of test was prioritized based on a wide range of patient characteristics including demographics, comorbid conditions, and severity of symptoms. Such targeted testing results in a highly selected group of patients who are tested for SARS-CoV-2.^21^ To adjust for unequal probabilities of being tested, we used the inverse probability weighting method, where the weight is based on the predicted probabilities (propensity score) of being tested, estimated by a logistic regression model using ADT exposure, selected patient covariates, and VHA facility.^22,23^ For this model, we implemented a nested case-control design with incidence density sampling to match each tested patient (case) to five patients who were eligible to be tested (controls) at the time of the case’s testing on age, race, and VHA facility.^24^

Generalized linear mixed models were used to determine associations between ADT exposure and severity of COVID-19 while controlling for patients’ demographic and clinical covariates, and the number of days from February 15 to the date of first SARS-CoV-2 positivity (as fixed effects) and VHA facility (as a random effect). All male Veterans in the study sample with a positive COVID-19 test were included in this analysis. We used SAS 9.2 (Cary, NC) for data preparation and all statistical analyses. Two-tailed hypothesis testing was performed using the significance level of 5%.

## RESULTS

### Patient characteristics

We identified 246,087 male patients tested for SARS-CoV-2 through July 15, 2020 (Table 1). Among the 246,087 tested patients, 3,057 patients were prescribed ADT and 243,030 were not. Compared to non-ADT patients, ADT patients were more likely to be Black (ADT 37%, no ADT 24%), older (mean age 73.9 vs 62.9; p<0.001), and had more comorbidities (Charlson Comorbidity Index ≥5 in 72% of ADT patients and 28% of non-ADT patients).

**Table 1.**
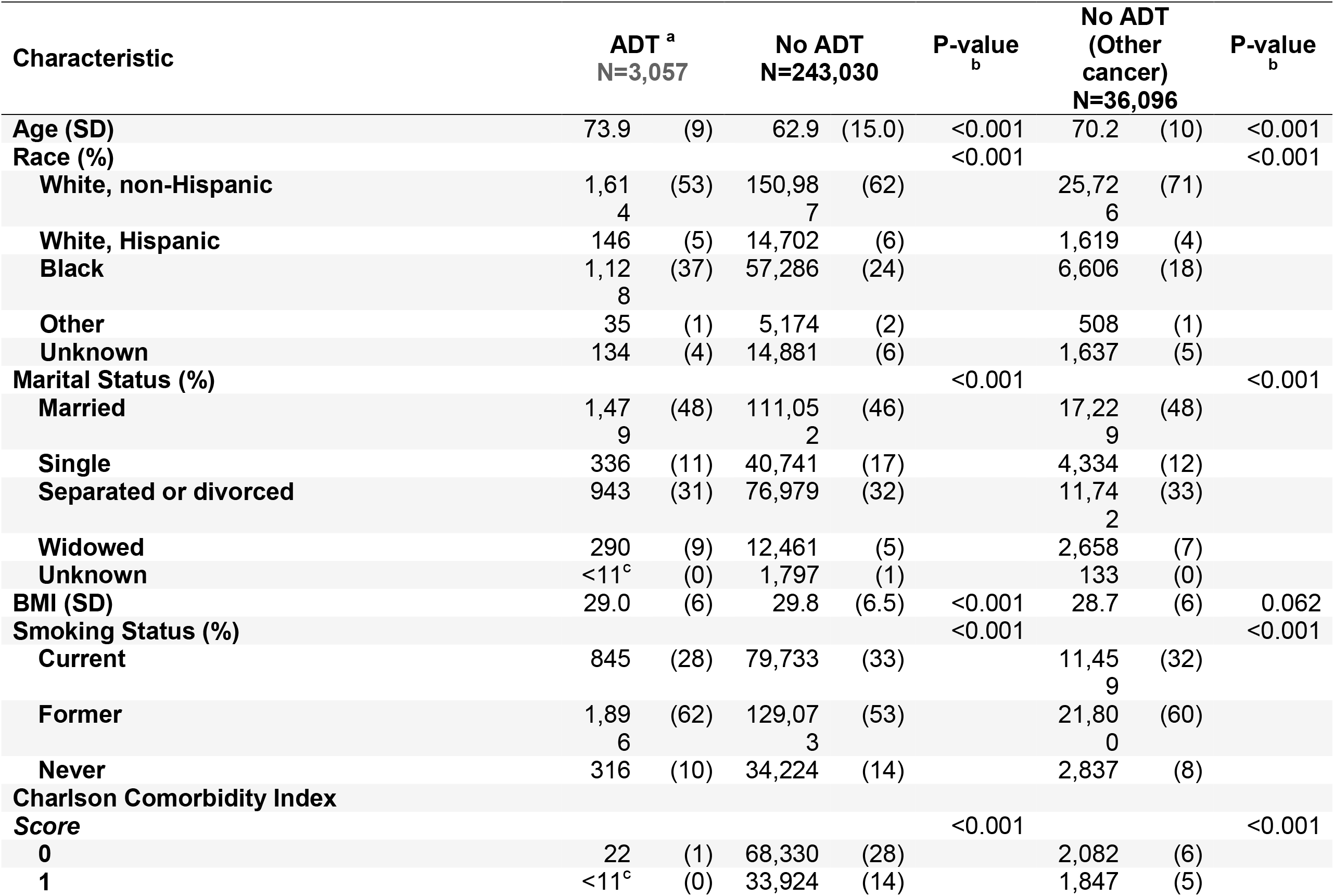
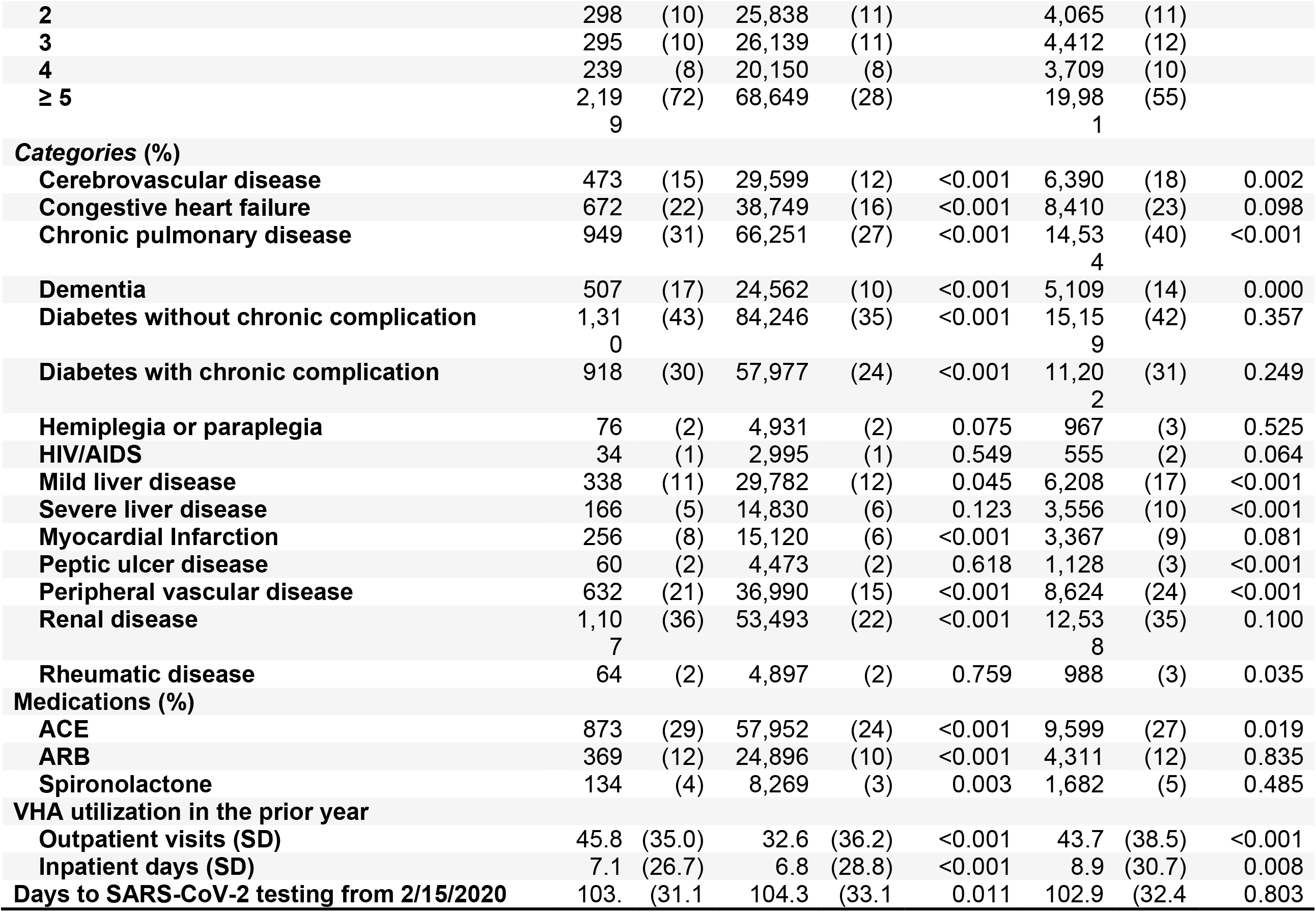
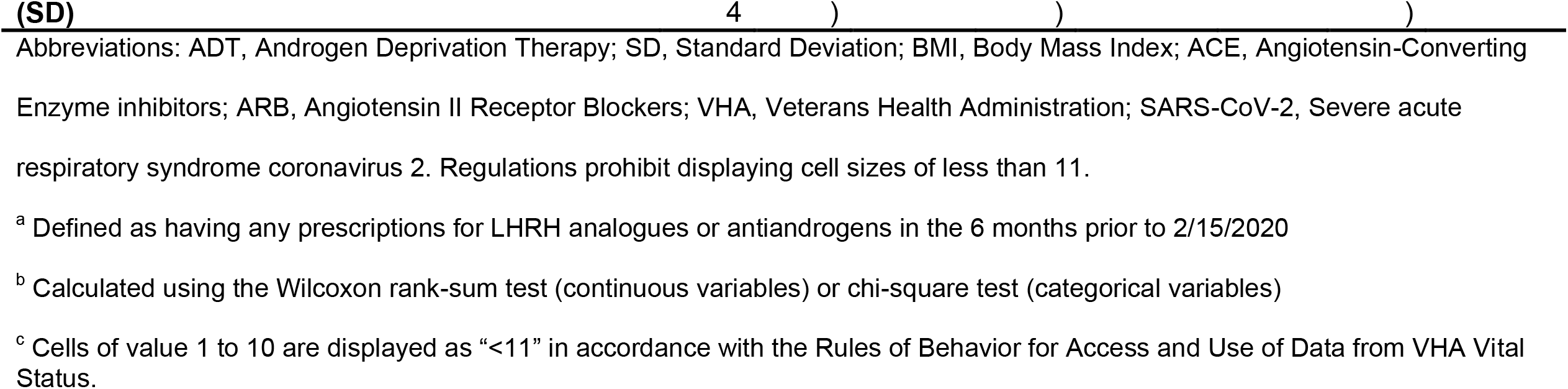
Baseline characteristics of patients tested for SARS-CoV-2

| Characteristic | ADT <sup>a</sup><br>N=3,057 |  | No ADT<br>N=243,030 |  | P-value<br><sub>b</sub> | No ADT<br>(Other cancer)<br>N=36,096 |  | P-value<br><sub>b</sub> |
| --- | --- | --- | --- | --- | --- | --- | --- | --- |
| **Age (SD)** | 73.9 | (9) | 62.9 | (15.0) | <0.001 | 70.2 | (10) | <0.001 |
| **Race (%)** |  |  |  |  | <0.001 |  |  | <0.001 |
| **White, non-Hispanic** | 1,614 | (53) | 150,987 | (62) |  | 25,726 | (71) |  |
| **White, Hispanic** | 146 | (5) | 14,702 | (6) |  | 1,619 | (4) |  |
| **Black** | 1,128 | (37) | 57,286 | (24) |  | 6,606 | (18) |  |
| **Other** | 35 | (1) | 5,174 | (2) |  | 508 | (1) |  |
| **Unknown** | 134 | (4) | 14,881 | (6) |  | 1,637 | (5) |  |
| **Marital Status (%)** |  |  |  |  | <0.001 |  |  | <0.001 |
| **Married** | 1,479 | (48) | 111,052 | (46) |  | 17,229 | (48) |  |
| **Single** | 336 | (11) | 40,741 | (17) |  | 4,334 | (12) |  |
| **Separated or divorced** | 943 | (31) | 76,979 | (32) |  | 11,742 | (33) |  |
| **Widowed** | 290 | (9) | 12,461 | (5) |  | 2,658 | (7) |  |
| **Unknown** | <11<sup>c</sup> | (0) | 1,797 | (1) |  | 133 | (0) |  |
| **BMI (SD)** | 29.0 | (6) | 29.8 | (6.5) | <0.001 | 28.7 | (6) | 0.062 |
| **Smoking Status (%)** |  |  |  |  | <0.001 |  |  | <0.001 |
| **Current** | 845 | (28) | 79,733 | (33) |  | 11,459 | (32) |  |
| **Former** | 1,896 | (62) | 129,073 | (53) |  | 21,800 | (60) |  |
| **Never** | 316 | (10) | 34,224 | (14) |  | 2,837 | (8) |  |
| **Charlson Comorbidity Index Score** |  |  |  |  | <0.001 |  |  | <0.001 |
| **0** | 22 | (1) | 68,330 | (28) |  | 2,082 | (6) |  |
| **1** | <11<sup>c</sup> | (0) | 33,924 | (14) |  | 1,847 | (5) |  |
| **2** | 298 | (10) | 25,838 | (11) |  | 4,065 | (11) |  |
| **3** | 295 | (10) | 26,139 | (11) |  | 4,412 | (12) |  |
| **4** | 239 | (8) | 20,150 | (8) |  | 3,709 | (10) |  |
| **≥ 5** | 2,199 | (72) | 68,649 | (28) |  | 19,981 | (55) |  |
| **Categories (%)** |  |  |  |  |  |  |  |  |
| **Cerebrovascular disease** | 473 | (15) | 29,599 | (12) | <0.001 | 6,390 | (18) | 0.002 |
| **Congestive heart failure** | 672 | (22) | 38,749 | (16) | <0.001 | 8,410 | (23) | 0.098 |
| **Chronic pulmonary disease** | 949 | (31) | 66,251 | (27) | <0.001 | 14,534 | (40) | <0.001 |
| **Dementia** | 507 | (17) | 24,562 | (10) | <0.001 | 5,109 | (14) | 0.000 |
| **Diabetes without chronic complication** | 1,310 | (43) | 84,246 | (35) | <0.001 | 15,159 | (42) | 0.357 |
| **Diabetes with chronic complication** | 918 | (30) | 57,977 | (24) | <0.001 | 11,202 | (31) | 0.249 |
| **Hemiplegia or paraplegia** | 76 | (2) | 4,931 | (2) | 0.075 | 967 | (3) | 0.525 |
| **HIV/AIDS** | 34 | (1) | 2,995 | (1) | 0.549 | 555 | (2) | 0.064 |
| **Mild liver disease** | 338 | (11) | 29,782 | (12) | 0.045 | 6,208 | (17) | <0.001 |
| **Severe liver disease** | 166 | (5) | 14,830 | (6) | 0.123 | 3,556 | (10) | <0.001 |
| **Myocardial Infarction** | 256 | (8) | 15,120 | (6) | <0.001 | 3,367 | (9) | 0.081 |
| **Peptic ulcer disease** | 60 | (2) | 4,473 | (2) | 0.618 | 1,128 | (3) | <0.001 |
| **Peripheral vascular disease** | 632 | (21) | 36,990 | (15) | <0.001 | 8,624 | (24) | <0.001 |
| **Renal disease** | 1,107 | (36) | 53,493 | (22) | <0.001 | 12,538 | (35) | 0.100 |
| **Rheumatic disease** | 64 | (2) | 4,897 | (2) | 0.759 | 988 | (3) | 0.035 |
| **Medications (%)** |  |  |  |  |  |  |  |  |
| **ACE** | 873 | (29) | 57,952 | (24) | <0.001 | 9,599 | (27) | 0.019 |
| **ARB** | 369 | (12) | 24,896 | (10) | <0.001 | 4,311 | (12) | 0.835 |
| **Spironolactone** | 134 | (4) | 8,269 | (3) | 0.003 | 1,682 | (5) | 0.485 |
| **VHA utilization in the prior year** |  |  |  |  |  |  |  |  |
| **Outpatient visits (SD)** | 45.8 | (35.0) | 32.6 | (36.2) | <0.001 | 43.7 | (38.5) | <0.001 |
| **Inpatient days (SD)** | 7.1 | (26.7) | 6.8 | (28.8) | <0.001 | 8.9 | (30.7) | 0.008 |
| **Days to SARS-CoV-2 testing from 2/15/2020** | 103. | (31.1) | 104.3 | (33.1) | 0.011 | 102.9 | (32.4) | 0.803 |

Abbreviations: ADT, Androgen Deprivation Therapy; SD, Standard Deviation; BMI, Body Mass Index; ACE, Angiotensin-Converting Enzyme inhibitors; ARB, Angiotensin II Receptor Blockers; VHA, Veterans Health Administration; SARS-CoV-2, Severe acute respiratory syndrome coronavirus 2. Regulations prohibit displaying cell sizes of less than 11.
<sup>a</sup> Defined as having any prescriptions for LHRH analogues or antiandrogens in the 6 months prior to 2/15/2020
<sup>b</sup> Calculated using the Wilcoxon rank-sum test (continuous variables) or chi-square test (categorical variables)
<sup>c</sup> Cells of value 1 to 10 are displayed as “<11” in accordance with the Rules of Behavior for Access and Use of Data from VHA Vital Status.

### SARS-CoV-2 positivity

The strategy for selecting patients to be included in the SARS-CoV-2 testing propensity score model is shown in Figure 1, and the baseline characteristics of all SARS-CoV-2-tested patients (cases) and the matched controls are shown in Table S1 in the Supplementary Appendix. ADT patients were more likely to be tested for SARS-CoV-2 (adjusted odds ratio (OR), 1.59 [95% confidence interval (CI), 1.52–1.68]; Table S2 in the Supplementary Appendix). In the primary, propensity-weighted comparison of ADT versus cancer patients not on ADT, ADT was associated with decreased likelihood of testing positive for SARS-CoV-2 after adjusting for the covariates listed in Table 1 (OR, 0.88 [95% CI, 0.81–0.95]; p=0.001; Table 2). Similar results were observed in secondary comparisons of ADT patients versus all non-ADT patients (OR, 0.75 [95% CI, 0.70–0.81]; p<0.001; Table 2) and patients with prostate cancer on ADT versus patients with prostate cancer not on ADT (OR, 0.85 [95% CI, 0.77–0.94]; p=0.002; Table S3 in the Supplementary Appendix).

**Figure 1.**
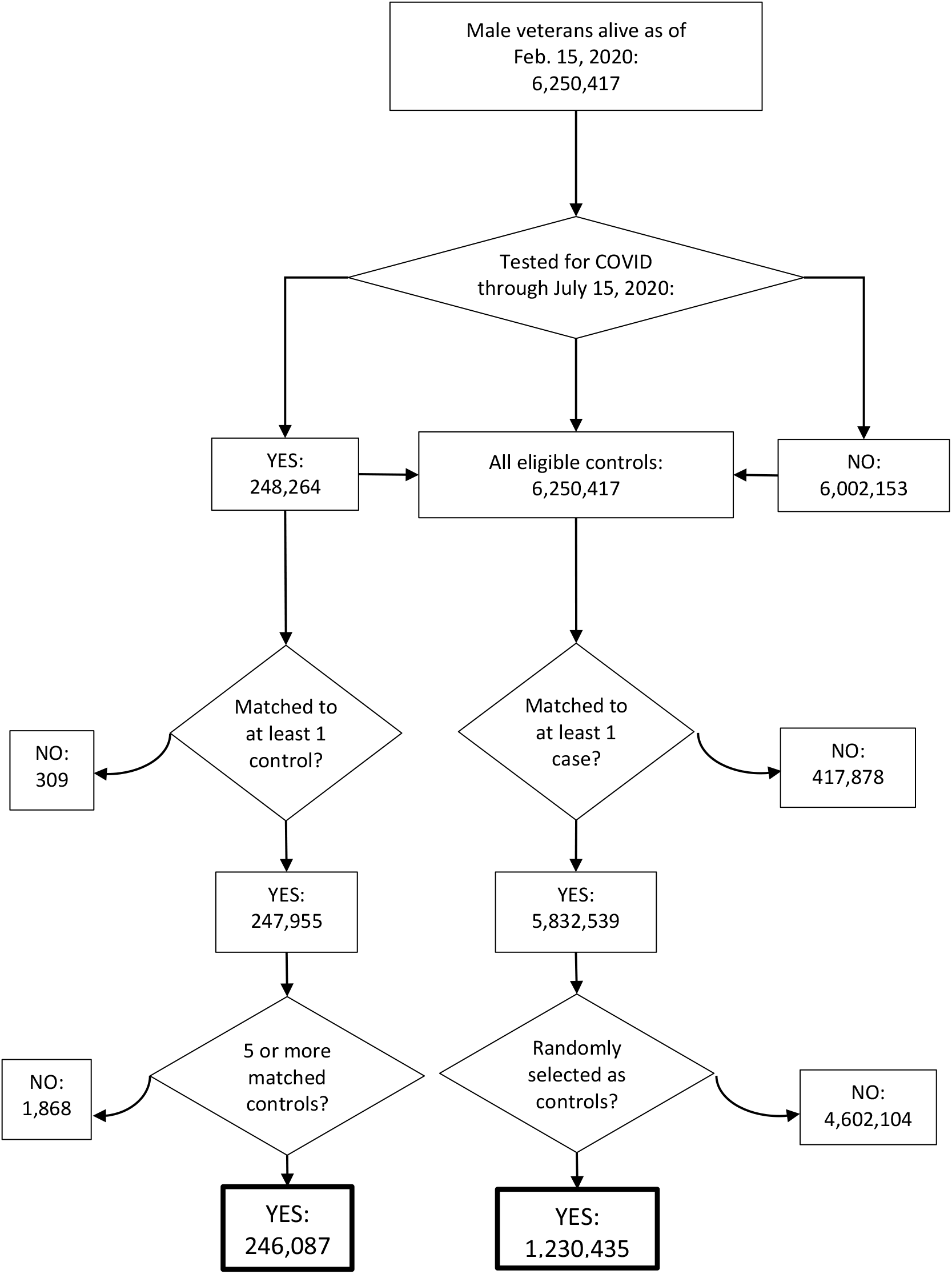
Consort diagram for the selection of patients for the model for the propensity to be tested for SARS-CoV-2. Curved arrows indicate excluded patients

**Table 2.**
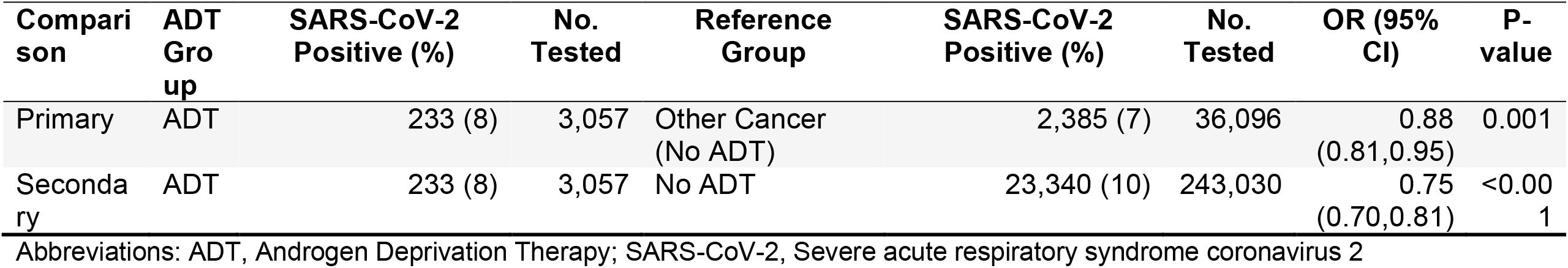
Association between ADT use and SARS-CoV-2 positivity

### COVID-19 severity

Our primary comparison of COVID-19 severity included 295 ADT patients with COVID-19 (including 189 with prostate cancer only and 97 with prostate cancer and a second cancer diagnosis), and 2,427 non-ADT, other cancer patients with COVID-19. Significant differences between the two groups included age, race, smoking status, and comorbidities (Table 3). Severe COVID-19 outcomes were observed in 76/295 (25%) of the ADT group and 727/2427 (30%) of the non-ADT group (Table 4). After controlling for clinical and demographic factors, ADT was associated with fewer severe COVID-19 outcomes (OR 0.72 [95% CI 0.53–0.96]; p=0.03; Table 4). Comparisons between ADT patients versus all patients not on ADT and prostate cancer patients on ADT versus prostate cancer patients not on ADT did not demonstrate a statistically significant protective effect of ADT (Table 4 and Table S3 in the Supplementary Appendix).

**Table 3.** Baseline characteristics at first positive SARS-CoV-2 test

| Characteristic | ADT <sup>a</sup><br>N=295 |  | No ADT<br>N=25,006 |  | P-value <sup>b</sup> | No ADT<br>(Other cancer)<br>N=2,427 |  | P-value <sup>b</sup> |
| --- | --- | --- | --- | --- | --- | --- | --- | --- |
| **Age (SD)** | 74.2 | (10.2) | 62.1 | (16.4) | <0.001 | 71.0 | (11.8) | <0.001 |
| **Race (%)** |  |  |  |  | <0.001 |  |  | <0.001 |
| White, non-Hispanic | 111 | (38) | 11,619 | (46) |  | 1,386 | (57) |  |
| White, Hispanic | 13 | (4) | 2,406 | (10) |  | 150 | (6) |  |
| Black | 152 | (52) | 8,580 | (34) |  | 752 | (31) |  |
| Other | <11<sup>d</sup> | (0) | 659 | (3) |  | 33 | (1) |  |
| Unknown | 18 | (6) | 1,742 | (7) |  | 106 | (4) |  |
| **Marital Status (%)** |  |  |  |  | 0.007 |  |  | 0.484 |
| Married | 141 | (48) | 11,686 | (47) |  | 1,134 | (47) |  |
| Single | 43 | (15) | 4,460 | (18) |  | 316 | (13) |  |
| Separated or divorced | 84 | (28) | 7,321 | (29) |  | 761 | (31) |  |
| Widowed | 27 | (9) | 1,278 | (5) |  | 200 | (8) |  |
| Unknown | 0 | (0) | 261 | (1) |  | 16 | (1) |  |
| **BMI (SD)** | 28.8 | (6.2) | 30.4 | (6.5) | <0.001 | 28.7 | (6.5) | 0.617 |
| **Smoking Status (%)** |  |  |  |  | 0.003 |  |  | 0.326 |
| Current | 71 | (24) | 5,927 | (24) |  | 650 | (27) |  |
| Former | 191 | (65) | 14,386 | (58) |  | 1,561 | (64) |  |
| Never | 33 | (11) | 4,693 | (19) |  | 216 | (9) |  |
| **Charlson Comorbidity Index Score** |  |  |  |  | <0.001 |  |  | <0.001 |
| 0 | <11<sup>d</sup> | (0) | 8,004 | (32) |  | 175 | (7) |  |
| 1 | <11<sup>d</sup> | (0) | 3,868 | (15) |  | 147 | (6) |  |
| 2 | 33 | (11) | 2,597 | (10) |  | 277 | (11) |  |
| 3 | 25 | (8) | 2,847 | (11) |  | 301 | (12) |  |
| 4 | 27 | (9) | 1,876 | (8) |  | 254 | (10) |  |
| ≥ 5 | 208 | (71) | 5,814 | (23) |  | 1,273 | (52) |  |
| **Categories (%)** |  |  |  |  |  |  |  |  |
| Cerebrovascular disease | 62 | (21) | 2,815 | (11) | <0.001 | 447 | (18) | 0.280 |
| Congestive heart failure | 77 | (26) | 3,314 | (13) | <0.001 | 576 | (24) | 0.368 |
| Chronic pulmonary disease | 91 | (31) | 5,160 | (21) | <0.001 | 843 | (35) | 0.184 |
| Dementia | 57 | (19) | 3,212 | (13) | 0.001 | 539 | (22) | 0.258 |
| Diabetes without chronic complication | 133 | (45) | 9,084 | (36) | 0.002 | 1,085 | (45) | 0.902 |
| Diabetes with chronic complication | 92 | (31) | 5,890 | (24) | 0.002 | 804 | (33) | 0.503 |
| Hemiplegia or paraplegia | <11<sup>d</sup> | (3) | 434 | (2) | 0.087 | 75 | (3) | 0.971 |
| HIV/AIDS | <11<sup>d</sup> | (1) | 333 | (1) | 0.800<sup>c</sup> | 43 | (2) | 0.605 |
| Mild liver disease | 33 | (11) | 2,369 | (9) | 0.318 | 370 | (15) | 0.064 |
| Severe liver disease | 22 | (7) | 1,092 | (4) | 0.010 | 201 | (8) | 0.626 |
| Myocardial Infarction | 24 | (8) | 1,088 | (4) | 0.002 | 199 | (8) | 0.970 |
| Peptic ulcer disease | 8 | (3) | 295 | (1) | 0.026<sup>c</sup> | 63 | (3) | 0.906 |
| Peripheral vascular disease | 62 | (21) | 3,040 | (12) | <0.001 | 527 | (22) | 0.784 |
| Renal disease | 110 | (37) | 4,987 | (20) | <0.001 | 844 | (35) | 0.393 |
| Rheumatic disease | <11<sup>d</sup> | (1) | 375 | (1) | 0.335<sup>c</sup> | 49 | (2) | 0.109 |
| **Medications (%)** |  |  |  |  |  |  |  |  |
| ACE | 90 | (27) | 5,134 | (21) | 0.005 | 529 | (22) | 0.038 |
| ARB | 39 | (13) | 2,489 | (10) | 0.063 | 296 | (12) | 0.613 |
| Spironolactone | <11<sup>d</sup> | (3) | 618 | (2) | 0.525 | 89 | (4) | 0.592 |
| **VHA utilization in the prior year** |  |  |  |  |  |  |  |  |
| Outpatient visits (SD) | 46.2 | (42.0) | 26.9 | (34.7) | <0.001 | 42.1 | (43.4) | <0.001 |
| Inpatient days (SD) | 8.2 | (29.9) | 7.0 | (35.7) | <0.001 | 14.2 | (50.9) | 0.668 |
| **Days to SARS-CoV-2 positivity from 2/15/2020 (SD)** | **96.2** | **(38.9)** | **101.0** | **(37.7)** | **0.022** | **95.6** | **(37.2)** | **0.972** |

Abbreviations: ADT, Androgen Deprivation Therapy; SD, Standard Deviation; BMI, Body Mass Index; CCI, Charlson Comorbidity
Index; ACE, Angiotensin-converting enzyme inhibitors; ARB, Angiotensin II Receptor Blockers; SARS-CoV-2, Severe acute respiratory syndrome coronavirus 2. Regulations prohibit displaying cell sizes of less than 11.
<sup>a</sup> Defined as having any prescriptions for luteinizing hormone releasing hormone analogues or antiandrogens in the 6 months prior to the date of first positive SARS-CoV-2 test.
<sup>b</sup> Calculated using the Wilcoxon rank-sum test (continuous variables) or chi-square test (categorical variables).
<sup>c</sup> Calculated using the two-sided Fisher's exact test.
<sup>d</sup> Cells of value 1 to 10 are displayed as "<11" in accordance with the Rules of Behavior for Access and Use of Data from VHA Vital Status.

**Table 4.**
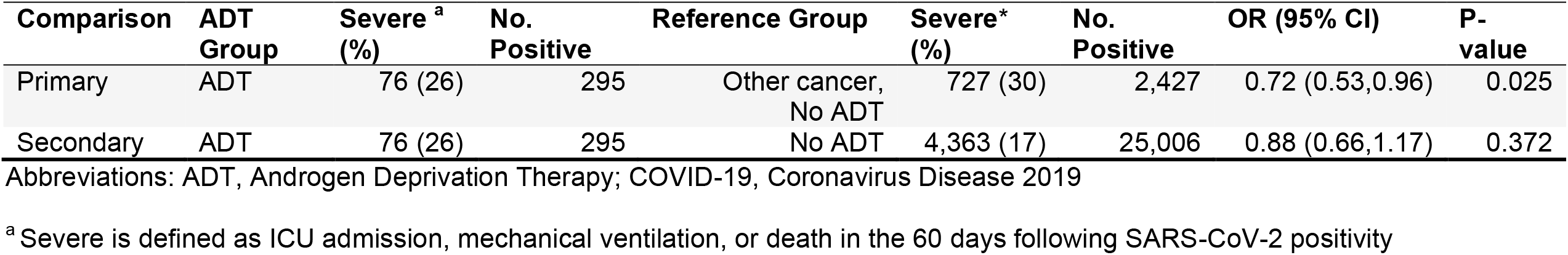
Association between ADT use and COVID-19 severity

## DISCUSSION

In our analysis of male U.S. Veterans tested for SARS-CoV-2, ADT use was associated with a reduced incidence and severity of COVID-19 compared to Veterans with non-prostate cancers who were not on ADT. The reduction in the incidence of COVID-19 was modest (OR 0.88) after correcting for baseline characteristics (e.g. smoking, cardiovascular disease and other comorbidities, and race) and the likelihood for undergoing SARS-CoV-2 testing. A reduced incidence of COVID-19 was also observed among ADT patients compared to all SARS-CoV-2-tested Veterans not on ADT as well as Veterans with prostate cancer not on ADT.

The effects of ADT on the incidence of COVID-19 has been reported by two groups. Montopoli et al. (2020) evaluated SARS-CoV-2 positive patients in the Veneto region of Italy and reported that patients with a cancer diagnosis other than prostate cancer as well as patients with prostate cancer not on ADT were over-represented compared to ADT patients (OR 4.86 [95% CI 1.88–12.56]; p=0.001, and OR 4.05 [95% CI 1.55–10.59]; p=0.004, respectively).

These results were based on COVID-19 incidence among all patients with cancer in the Veneto region, irrespective of whether these patients had been tested for SARS-CoV-2. Whereas the results from the Italian study are consistent with the protective effect of ADT on COVID-19 incidence in the analogous populations in the Veteran population, the magnitude of the effect was far greater in the Veneto population. In contrast, Klein et al. (2020), reporting on a cohort of patients from an Ohio healthcare system, observed no protective effect of ADT on incidence of COVID-19 in prostate cancer patients on ADT versus prostate cancer patients not on ADT.

The association of ADT with the severity of COVID-19 has been reported in three studies. In the Italian study, ADT was associated with a reduced likelihood of severe disease (ICU admission and death), but this was premised on only four COVID-19 positive patients on ADT (only 1 of whom developed severe disease), 13 COVID-19 positive patients with prostate cancer not on ADT, and 89 COVID-19 positive patients with other cancers not on ADT.^17^ By contrast, the Ohio study did not identify a protective effect of ADT on the severity of COVID-19 outcomes, but the authors recognized that robust statistics were not feasible due to limited sample size, despite being significantly larger than the Italian cohort.^18^ Based on a small cohort of ADT and non-ADT patients (n=22 and 36, respectively), Patel et al. (2020) reported that ADT was associated with a reduced risk of hospitalization (OR 0.23 [95% CI 0.06–0.79]; p<0.02) and supplemental oxygen use (OR 0.26 [95% CI 0.07–0.92]; p=0.04), and a non-statistically significant trend toward a reduction in mechanical ventilation and death after correcting for age, cardiac disease, and pulmonary disease.^25^ By comparison, our study demonstrated ADT exposure was associated with a reduction in COVID-19 severity (OR 0.72) compared to cancer patients without ADT exposure, based on a composite endpoint of ICU admission, mechanical ventilation, and death.

Strengths of this study include a multivariate regression analysis to account for differences in baseline characteristics, which was not performed in the Italian study, as well as a much larger cohort of patients in comparison to the Italian study, though still an order of magnitude smaller than the current VHA cohort (e.g. n=304 and n=3,057 patients on ADT in the Ohio and Veteran cohorts, respectively). Importantly, neither the Italian nor the Ohio study accounted for differences in the likelihood to be tested for SARS-CoV-2 among the respective populations, which can bias the COVID-19 incidence results, especially if ADT patients are more likely to be tested for SARS-CoV-2. Indeed, in our Veteran population, the adjusted odds ratio for SARS-CoV-2 testing was 1.59 for ADT patients, which we incorporated as a propensity score in our analyses of COVID-19 incidence. In addition, our study is less subject to ascertainment bias, since Veterans largely seek care within the VA healthcare system. Weaknesses include the retrospective nature of the analysis and uncertainty about the underlying cause for ICU admission, mechanical ventilation, and death. While we treated VHA facility as a random effect in our severity analysis, the rapid change in treatment decisions and ability for hospitals to handle capacity at different times in the pandemic may leave confounding for patients who develop severe disease.

Ongoing work in VA may provide further clarify the association between ADT and COVID-19 illness and determine the utility of ADT agents including LHRH analogs, androgen receptor antagonists, and 5-alpha reductase inhibitors. LHRH analogs achieve castrate levels of serum testosterone in virtually all patients, although the LHRH antagonist, degarelix, is the only FDA-approved LHRH analog that rapidly suppresses serum testosterone concentrations. A randomized, double-blind, placebo-controlled, multicenter VA trial entitled, “Hormonal Intervention for the Treatment of Veterans with COVID-19 Requiring Hospitalization (HITCH)” (Clinicaltrials.gov identifier NCT04397718), is prospectively testing the hypothesis that ADT with the LHRH antagonist, degarelix, reduces the severity of COVID-19 illness defined by a composite of ongoing hospitalization, intubation, or death within 15 days of randomization. The VA Million Veteran Program MVP035 COVID-19 Disease Mechanisms study is investigating the role of TMPRSS2 and other androgen-regulated genes in COVID-19 severity. Several other ongoing trials in the US and abroad are investigating the effects of androgen receptor antagonists (e.g. bicalutamide, enzalutamide) on COVID-19 illness.^26^

There is a paucity of effective treatments for severe COVID-19. In the Solidarity trial, none of remdesivir, lopinavir, hydroxychloroquine or interferon beta-1a affected mortality, initiation of ventilation or duration of hospitalization, although the ACT-1 study demonstrated that remdesivir reduces duration of hospitalization.^27,28^ Convalescent plasma, despite its initial promise, failed to improve severity or mortality of hospitalized COVID-19 patients; likewise, IL-6 pathway inhibitors have not improved survival of severely ill patients.^29^ In contrast, dexamethasone is the only treatment that has been consistently shown in randomized controlled trials to improve survival and risk of intubation of COVID-19 patients.^30,31^

## CONCLUSIONS

Our analysis of Veterans enrolled in the VHA suggests an association of ADT exposure with reduced incidence and severity of COVID-19. Repurposing of drugs that modulate androgen production and/or action may represent viable potential treatments for COVID-19.

## Data Availability

We used the Corporate Data Warehouse (CDW) of the VHA, a national data repository that provides access to the electronic health records of all individuals who received care in the VHA. In addition, we drew from the VA COVID-19 Shared Data Resource (CSDR), a newly developed data domain that consists of a wide range of information related to COVID-19 available for all patients who received a COVID-19 laboratory test within VHA or whose positive test result outside VHA was recorded in VHA clinical notes.

## Notes

Funding: Funding for this study was provided by the US Department of Veterans Affairs (VA), Veterans Health Administration, Cooperative Studies Program, grant number 825-MS-DI-33848, and used resources and facilities at the VA Informatics and Computing Infrastructure (VINCI), VA HSR RES 13-457. Dr. Rettig is supported by The Prostate Cancer Foundation (PCF) David Geffen Precision Oncology Center of Excellence of VAGLAHS and a Department of Veterans Affairs Merit Review Award. Drs. Becker and Makarov are supported by The John and Daria Barry Precision Oncology Center of Excellence of the VANYHHS, and Edward Blank and Sharon Cosloy–Blank Family Foundation. Dr. Becker is supported by VA Career Development Award (CDA 16-206). Dr. Makarov is a Prostate Cancer Foundation Young Investigator Awardee.

### Competing Interest Statement

Disclosures: SL reports equity in Gilead Sciences, Inc. SLD reports research grants from the following for-profit organizations outside this submitted work: Alnylam Pharmaceuticals Inc., AbbVie Inc., Astellas Pharma Inc., AstraZeneca Pharmaceuticals LP, Biodesix, Inc., Boehringer Ingelheim International GmbH, Celgene Corporation, Eli Lilly and Company, Genentech Inc., Gilead Sciences Inc., GlaxoSmithKline PLC, Innocrin Pharmaceuticals Inc., Janssen Pharmaceuticals, Inc., Kantar Health, Myriad Genetic Laboratories, Inc., Novartis International AG, and Parexel International Corporation through the University of Utah or Western Institute for Veteran Research.

### Funding Statement

Funding: Funding for this study was provided by the US Department of Veterans Affairs (VA), Veterans Health Administration, Cooperative Studies Program, grant number 825 MS DI 33848, and used resources and facilities at the VA Informatics and Computing Infrastructure (VINCI), VA HSR RES 13 457. Dr. Rettig is supported by The Prostate Cancer Foundation (PCF) David Geffen Precision Oncology Center of Excellence of VAGLAHS and a Department of Veterans Affairs Merit Review Award. Drs. Becker and Makarov are supported by The John and Daria Barry Precision Oncology Center of Excellence of the VANYHHS, and Edward Blank and Sharon Cosloy Blank Family Foundation. Dr. Becker is supported by VA Career Development Award (CDA 16 206). Dr. Makarov is a Prostate Cancer Foundation Young Investigator Awardee.

### Author Declarations

This study was approved by the VA Central Institutional Review Board.

