## Supplemental Table 1 for "A population-level analysis of the protective effects of androgen deprivation therapy against COVID-19 disease incidence and severity"

**Table S1**. Baseline characteristics of base sample and matched sample

|  | Base | | | | Matched | | | |
| --- | --- | --- | --- | --- | --- | --- | --- | --- |
|  | **Tested** | | **Not tested** | | **Case** | | **Control** | |
| Number of patients | 248,264 |  | 6,002,153 |  | 246,087 |  | 1,230,435 |  |
| ADT (%) | 3,088 | (1.2) | 25,224 | (0.4) | 3,057 | (1.2) | 6,272 | (0.5) |
| Age (years) (%) |  |  |  |  |  |  |  |  |
| Mean (SD) | 63.0 | (15.0) | 62.6 | (16.8) | 63.0 | (15.0) | 63.0 | (15.0) |
| < 45 | 34,831 | (14) | 1,101,250 | (18) | 34,241 | (14) | 171,205 | (14) |
| 45-54 | 27,986 | (11) | 688,992 | (11) | 27,677 | (11) | 138,385 | (11) |
| 55-64 | 53,955 | (22) | 982,064 | (16) | 53,616 | (22) | 268,080 | (22) |
| 65-74 | 83,748 | (34) | 1,829,250 | (30) | 83,463 | (34) | 417,315 | (34) |
| 75-84 | 33,005 | (13) | 908,966 | (15) | 32,757 | (13) | 163,785 | (13) |
| ≥ 85 | 14,739 | (6) | 491,631 | (8) | 14,333 | (6) | 71,665 | (6) |
| Race (%) |  |  |  |  |  |  |  |  |
| White, non-Hispanic | 152,651 | (61) | 4,016,395 | (67) | 152,601 | (62) | 763,005 | (62) |
| White, Hispanic | 15,260 | (6) | 284,011 | (5) | 14,848 | (6) | 74,240 | (6) |
| Black | 58,701 | (24) | 919,061 | (15) | 58,414 | (24) | 292,070 | (24) |
| HI/PI | 2,034 | (1) | 49,117 | (1) | 1,520 | (1) | 7,600 | (1) |
| AI/AK | 1,993 | (1) | 45,026 | (1) | 1,488 | (1) | 7,440 | (1) |
| Asian | 2,543 | (1) | 64,830 | (1) | 2,201 | (1) | 11,005 | (1) |
| Unknown | 15,082 | (6) | 623,713 | (10) | 15,015 | (6) | 75,075 | (6) |
| Marital status (%) |  |  |  |  |  |  |  |  |
| Married | 113,540 | (46) | 3,397,095 | (57) | 112,531 | (46) | 669,013 | (54) |
| Single | 41,480 | (17) | 874,191 | (15) | 41,077 | (17) | 182,585 | (15) |
| Separated or divorced | 78,488 | (32) | 1,365,029 | (23) | 77,922 | (32) | 312,150 | (25) |
| Widowed | 12,927 | (5) | 251,705 | (4) | 12,751 | (5) | 48,705 | (4) |
| Unknown | 1,829 | (1) | 114,133 | (2) | 1,806 | (1) | 17,982 | (1) |
| BMI (%) |  |  |  |  |  |  |  |  |
| Mean (SD) | 29.8 | (6.4) | 29.7 | (5.8) | 29.8 | (6.4) | 29.8 | (5.9) |
| < 18.5 | 3,992 | (2) | 48,488 | (1) | 3,958 | (2) | 10,747 | (1) |
| 18.5-24.9 | 51,195 | (21) | 1,100,654 | (18) | 50,675 | (21) | 227,021 | (18) |
| 25.0-29.9 | 83,237 | (34) | 2,169,578 | (36) | 82,520 | (34) | 443,159 | (36) |
| ≥ 30 | 108,494 | (44) | 2,493,168 | (42) | 107,609 | (44) | 521,060 | (42) |
| Unknown | 1,346 | (1) | 190,265 | (3) | 1,325 | (1) | 28,448 | (2) |
| Smoking status (%) |  |  |  |  |  |  |  |  |
| Current | 81,227 | (33) | 1,619,508 | (27) | 80,578 | (33) | 350,277 | (28) |
| Former | 132,108 | (53) | 2,970,410 | (49) | 130,969 | (53) | 608,266 | (49) |
| Never | 34,929 | (14) | 1,412,235 | (24) | 34,540 | (14) | 271,892 | (22) |
| CCI Categories (%) |  |  |  |  |  |  |  |  |
| Cerebrovascular disease | 30,288 | (12) | 307,669 | (5) | 30,072 | (12) | 69,586 | (6) |
| Congestive heart failure | 39,713 | (16) | 307,897 | (5) | 39,421 | (16) | 71,017 | (6) |
| Chronic pulmonary disease | 67,640 | (27) | 758,051 | (13) | 67,200 | (27) | 166,445 | (14) |
| Dementia | 25,365 | (10) | 192,304 | (3) | 25,069 | (10) | 40,135 | (3) |
| Diabetes without chronic complication | 86,249 | (35) | 1,304,921 | (22) | 85,556 | (35) | 295,000 | (24) |
| Diabetes with chronic complication | 59,354 | (24) | 697,034 | (12) | 58,895 | (24) | 160,550 | (13) |
| Hemiplegia or paraplegia | 5,032 | (2) | 28,439 | (0) | 5,007 | (2) | 7,436 | (1) |
| HIV/AIDS | 3,048 | (1) | 22,628 | (0) | 3,029 | (1) | 6,966 | (1) |
| Mild liver disease | 30,345 | (12) | 269,707 | (4) | 30,120 | (12) | 69,213 | (6) |
| Severe liver disease | 15,095 | (6) | 110,360 | (2) | 14,996 | (6) | 31,710 | (3) |
| Localized solid tumor | 53,487 | (22) | 666,305 | (11) | 53,155 | (22) | 150,531 | (12) |
| Metastatic solid tumor | 6,115 | (2) | 30,613 | (1) | 6,078 | (2) | 7,609 | (1) |
| Myocardial Infarction | 15,474 | (6) | 104,337 | (2) | 15,376 | (6) | 25,004 | (2) |
| Peptic ulcer disease | 4,569 | (2) | 31,019 | (1) | 4,533 | (2) | 7,681 | (1) |
| Peripheral vascular disease | 37,885 | (15) | 363,926 | (6) | 37,622 | (15) | 81,429 | (7) |
| Renal disease | 55,037 | (22) | 541,912 | (9) | 54,600 | (22) | 122,707 | (10) |
| Rheumatic disease | 4,994 | (2) | 64,291 | (1) | 4,961 | (2) | 13,796 | (1) |
| Medications (%) |  |  |  |  |  |  |  |  |
| ACE | 59,233 | (24) | 960,899 | (16) | 58,825 | (24) | 219,513 | (18) |
| ARB | 25,454 | (10) | 384,629 | (6) | 25,265 | (10) | 86,733 | (7) |
| Spironolactone | 8,447 | (3) | 78,357 | (1) | 8,403 | (3) | 18,440 | (1) |
| VHA utilization in the prior year |  |  |  |  |  |  |  |  |
| Outpatient visits (SD) | 32.7 | (36.2) | 10.6 | (15.5) | 32.7 | (36.2) | 12.5 | (17.8) |
| Inpatient days (SD) | 6.9 | (28.8) | 0.7 | (7.2) | 6.9 | (28.7) | 1.0 | (8.5) |

Abbreviations: ADT, Androgen Deprivation Therapy; SD, Standard Deviation; BMI, Body Mass Index; CCI, Charlson Comorbidity Index; ACE, Angiotensin-converting enzyme inhibitors; ARB, Angiotensin II Receptor Blockers; VHA, Veterans Health Administration.
