## Supplemental Table 2 for "A population-level analysis of the protective effects of androgen deprivation therapy against COVID-19 disease incidence and severity"

**Table S2**. Logistic regression results of propensity score model to predict receipt of SARS-CoV-2 testing

|  | Unadjusted | | | Fully adjusted | |
| --- | --- | --- | --- | --- | --- |
|  | **OR (95% CI)** | | | **OR (95% CI)** | |
| ADT | 2.46 (2.35,2.56) | | | 1.59 | (1.52,1.67) |
| Age |  |  |  | 0.99 | (0.99,0.99) |
| Race |  |  |  |  |  |
| White, non-Hispanic (Ref). |  |  |  |  |  |
| White, Hispanic |  |  |  | 0.96 | (0.94,0.98) |
| Black |  |  |  | 0.79 | (0.78,0.80) |
| Other |  |  |  | 1.00 | (0.97,1.03) |
| Unknown |  |  |  | 1.27 | (1.24,1.29) |
| Marital status |  |  |  |  |  |
| Married (Ref.) |  |  |  |  |  |
| Single |  |  |  | 1.22 | (1.20,1.24) |
| Separated or divorced |  |  |  | 1.27 | (1.26,1.28) |
| Widowed |  |  |  | 1.34 | (1.31,1.37) |
| Unknown |  |  |  | 0.72 | (0.68,0.76) |
| BMI |  |  |  | 0.99 | (0.99,1.00) |
| Smoking status |  |  |  |  |  |
| Current (Ref.) |  |  |  |  |  |
| Former |  |  |  | 1.01 | (1.00,1.02) |
| Never |  |  |  | 0.82 | (0.81,0.83) |
| Comorbidities |  |  |  |  |  |
| Cerebrovascular disease |  |  |  | 1.18 | (1.16,1.20) |
| Congestive heart failure |  |  |  | 1.30 | (1.28,1.32) |
| Chronic pulmonary disease |  |  |  | 1.45 | (1.44,1.47) |
| Dementia |  |  |  | 1.61 | (1.57,1.64) |
| Diabetes without chronic complication |  |  |  | 1.05 | (1.04,1.07) |
| Diabetes with chronic complication |  |  |  | 1.08 | (1.06,1.09) |
| Hemiplegia or paraplegia |  |  |  | 1.16 | (1.11,1.21) |
| HIV/AIDS |  |  |  | 1.40 | (1.34,1.47) |
| Mild liver disease |  |  |  | 1.35 | (1.33,1.38) |
| Severe liver disease |  |  |  | 0.95 | (0.92,0.98) |
| Myocardial Infarction |  |  |  | 1.22 | (1.19,1.25) |
| Peptic ulcer disease |  |  |  | 1.32 | (1.27,1.38) |
| Peripheral vascular disease |  |  |  | 1.23 | (1.21,1.25) |
| Renal disease |  |  |  | 1.30 | (1.29,1.32) |
| Rheumatic disease |  |  |  | 1.19 | (1.15,1.23) |
| Medications |  |  |  |  |  |
| ACE |  |  |  | 1.14 | (1.13,1.15) |
| ARB |  |  |  | 1.13 | (1.11,1.15) |
| Spironolactone |  |  |  | 1.00 | (0.97,1.03) |
| VHA utilization in the prior year |  |  |  |  |  |
| Outpatient visit |  |  |  | 1.03 | (1.03,1.03) |
| Inpatient days |  |  |  | 1.00 | (1.00,1.00) |

Abbreviations: ADT, Androgen Deprivation Therapy; SD, Standard Deviation; BMI, Body Mass Index; ACE, Angiotensin-converting enzyme inhibitors; ARB, Angiotensin II Receptor Blockers; VHA, Veterans Health Administration.
