## Supplemental Table 3 for "A population-level analysis of the protective effects of androgen deprivation therapy against COVID-19 disease incidence and severity"

**Table S3**. Association between ADT use and SARS-CoV-2 positivity and COVID-19 severity among prostate cancer patients

|  | ADT | | No ADT (reference) | |  |  |  |
| --- | --- | --- | --- | --- | --- | --- | --- |
| Outcome | **Event (%)** | **No. Tested/Positive** | **Event (%)** | **No. Tested/Positive** | **OR** | **(95% CI)** | **P-value** |
| SARS-CoV-2 positivity | 149 (8) | 1,908 | 1228 (10) | 12,741 | 0.85 | (0.77,0.94) | 0.002 |
| Severe COVID-19^*^ | 42 (22) | 189 | 316 (23) | 1,346 | 0.97 | (0.65,1.45) | 0.893 |

Abbreviations: ADT, Androgen Deprivation Therapy; SARS-CoV-2, Severe acute respiratory syndrome coronavirus 2; COVID-19, Coronavirus Disease 2019.

*Severe COVID-19 is defined as ICU admission, mechanical ventilation, or death in the 60 days following SARS-CoV-2 positivity.
